# The Importance of Automation in Genetic Diagnosis: Lessons from Analyzing an Inherited Retinal Degeneration Cohort with the Mendelian Analysis Toolkit (MATK)

**DOI:** 10.1101/2021.04.09.21255188

**Authors:** Erin Zampaglione, Matthew Maher, Emily M. Place, Naomi E. Wagner, Stephanie DiTroia, Katherine R. Chao, Eleina England, Broad CMG, Andrew Catomeris, Sherwin Nassiri, Seraphim Himes, Joey Pagliarulo, Charles Ferguson, Eglé Galdikaité-Braziené, Brian Cole, Eric A. Pierce, Kinga M. Bujakowska

**Author notes:** Corresponding author: Dr. Kinga Bujakowska, Massachusetts Eye and Ear, 243 Charles Street, Boston, MA 02114.

## Abstract

**Purpose:** In Mendelian disease diagnosis, variant analysis is a repetitive, error-prone, and time-consuming process. To address this, we have developed the Mendelian Analysis Toolkit (MATK), a configurable automated variant ranking program.

**Methods:** MATK aggregates variant information from multiple annotation sources and uses expert-designed rules with parameterized weights to produce a ranked list of potentially causal solutions. MATK performance was measured by a comparison of MATK-aided versus human domain-expert analyses of 1060 inherited retinal degeneration (IRD) families investigated with an IRD-specific gene panel (589 individuals) and exome sequencing (471 families).

**Results:** When comparing MATK-assisted analysis to expert curation in both IRD-specific and exome sequencing (1060 subjects), 97.3% of potential solutions found by experts were also identified by the MATK-assisted analysis (541 solutions identified with MATK of 556 solutions found by conventional analysis). Furthermore, MATK-assisted analysis identified 114 additional potential solutions from the 504 cases unsolved by the conventional analysis.

**Conclusion:** MATK expedites the process of identifying likely solving variants in Mendelian traits and reduces variability coming from human error and researcher bias. MATK facilitates data re-analysis to keep up with the constantly improving annotation sources and NGS processing pipelines. The software is open source, available at https://gitlab.com/matthew_maher/mendelanalysis

## INTRODUCTION

Next Generation Sequencing (NGS) has been increasingly used in the study of Mendelian diseases for both new gene discovery and clinical diagnosis^1^. However, as more sequencing data becomes available, the ability to analyze that data must scale appropriately^2^. Automation in read alignment and variant calling is a rapidly advancing field, which has shown consistent progress in speed and accuracy^3,4^. Similarly, the tools and datasets available for variant annotation are constantly improving and expanding^5,6^, as seen in the human genome reference, transcript information, variant population frequency^7,8^ and predicted variant effects^9^. However, the interpretation of variants and the determination of a genetic diagnosis remains a primarily manual and iterative task which requires analysts to integrate information from multiple sources. Analysis of single nucleotide variant (SNV) and structural variant (SV) calls generally involves hard filters to reduce the number of plausible causative variants and further cross-referencing with multiple online databases, such as OMIM, ClinVar^10^ and HGMD^11^ (Figure 1A). While guidelines have been developed to standardize both SNV and SV interpretation^12,13^ and some work has been done to automate the use of these guidelines^14^, the task remains tedious, error-prone, and highly dependent on the analysts’ experience. Furthermore, while the upstream pipelines of variant calling and annotation constantly improve, individual subject data is rarely revisited by the manual curators, even though variant interpretation has been shown to clearly benefit from periodic reanalysis^15–17^. The motivation for the development of the Mendelian Analysis Toolkit (MATK) was to increase automation, accuracy, and repeatability of this process.

**Figure 1:**
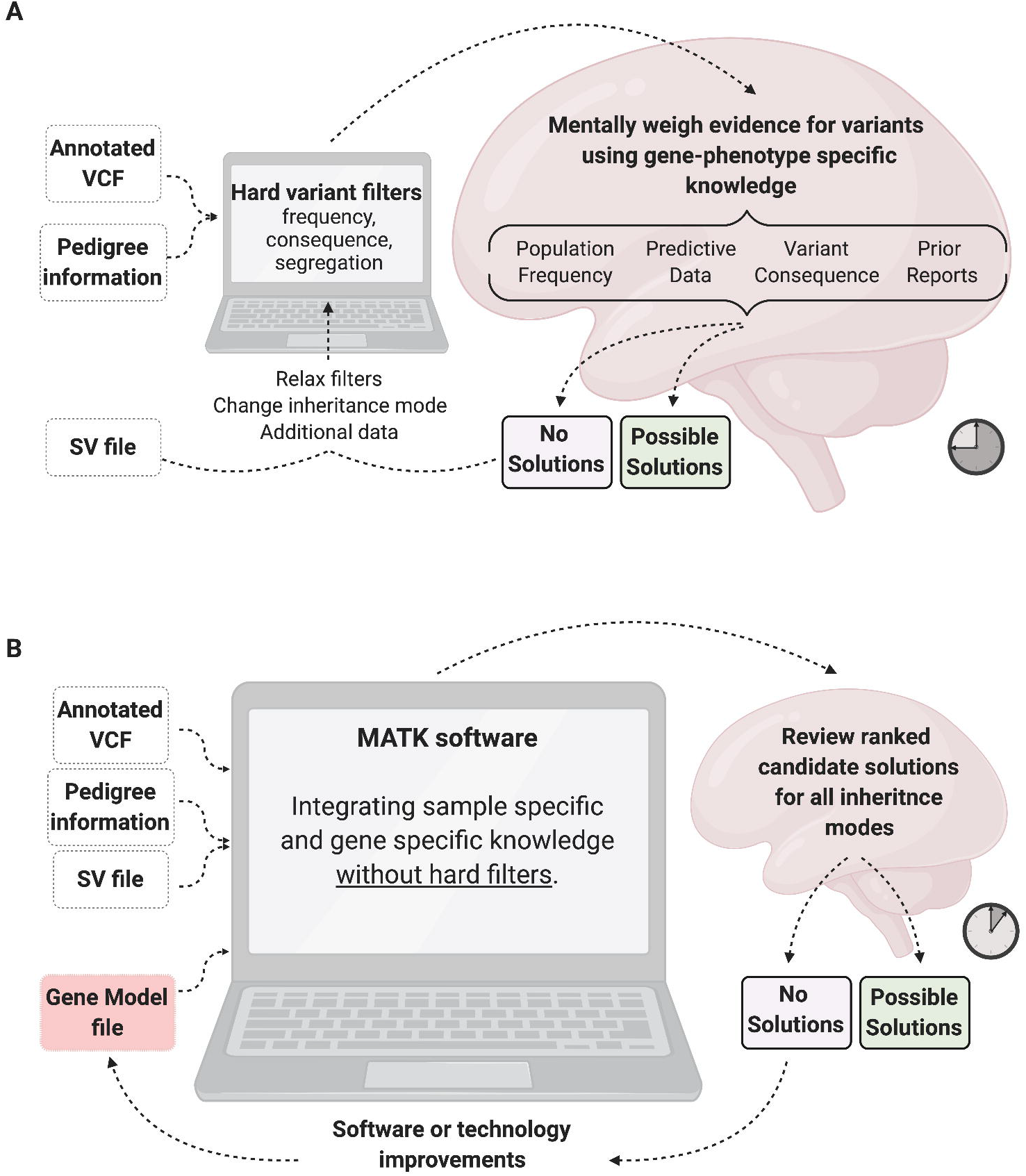
Standard variant assessment protocol for human analysts. A) The process of NGS variant analysis without MATK, which involves hard filtering of variants down to a manageable list, then using the analyst’s disease-specific expertise to weigh the evidence for each variant, often going through multiple iterations of filtering. B) In the analysis with MATK, much of the domain expertise is encoded in the Gene Model File (GMF) and the evidence is weighed by the Annotation Binding Code (ABC) which allows for a standardized analysis. The GMF and the ABC are fully customizable. Created with BioRender.com

MATK is a software suite designed to prioritize variants that may be causal for a Mendelian disease. The software is customizable, intended to be tailored to the annotation pipeline and evolving knowledge of genes relevant to the disease under study (Figure 1B). MATK can perform pedigree-aware analysis and incorporate other inputs such as Copy Number Variation (CNV) predictions or arbitrary gene-level annotations such as probability of Loss-of-Function Intolerance (pLI) scores^7^ and tissue-specific RNA expression data^18^.

We have validated MATK using a cohort of subjects with inherited retinal degeneration (IRD)^19^. This disease space is an excellent test case for MATK, as IRDs are highly heterogeneous, with over 270 causal genes, following all modes of Mendelian inheritance^20^. Previous work has shown that ∼60% of cases can be genetically solved by SNVs and/or CNVs in the exons of known IRD genes^21,22^. We show that MATK performs similarly to human analysts for both panel-based sequencing and exome sequencing (ES), while providing improvements in analysis efficiency, consistency and repeatability.

## METHODS

### Ethical guidelines

The study was approved by the Institutional Review Board at the Massachusetts Eye and Ear (Human Studies Committee MEE, Mass General Brigham, USA) and adhered to the tenets of the Declaration of Helsinki. Informed consent was obtained from all individuals on whom genetic testing and further molecular evaluations were performed.

#### Sequencing and Annotation

(extended version present in Supplemental Methods) Genetic Eye Disease (GEDi) sequencing was performed as described previously^21^. VCF files were generated using the Genome Analysis Toolkit (GATK) version 3 (https://software.broadinstitute.org/gatk/), and annotated using VEP^6^ and VCFAnno^23^. VEP provided transcript-specific sequence-consequence annotations from GENCODE v19 and regulatory annotations from ENCODE^24^. VCFAnno was used to annotate the variants with gnomAD^7,8^, ClinVar^10^, HGMD^11^, and CADD^9^. CNV predictions were produced using gCNV^4^, and the known *MAK*-Alu structural variant was identified using a custom script^25^.

Exome sequencing was performed at the Center for Mendelian Genomics at the Broad Institute of MIT and Harvard using methodology described previously^7^. Exome sequencing data was aligned to human genome 38. Variants were called using Genome Analysis Toolkit (GATK) HaplotypeCaller package version 3.5. The data were displayed and analyzed with an online tool (https://seqr.broadinstitute.org).

### MATK configuration

Variant-level information used in this study consisted of an annotated variant call format (VCF) file and CNV predictions from gCNV. For the GEDi panel sequence analyses, MATK’s Annotation Binding Code (ABC) was configured based on prior expert knowledge as a summation of 6 unique functions pertaining to 1) sequence consequence, 2) population frequency, 3) ClinVar pathogenicity classification, 4) HGMD pathogenicity classification, 5) CADD score, and 6) ENCODE promoter status, with the final score capped at 20 (Figure 2A, Supplementary Table 1A). For GEDi analyses, the gene-level information, contained in the Gene Model File (GMF), was configured with a default maximum likely frequency for autosomal recessive (AR) and X linked (XL) genes at 0.002, and for autosomal dominant (AD) genes at 0.00001, although exceptions were made for some genes based on known allele frequencies for IRD variants (Figure 2A, Supplementary Table 2). Once MATK had ranked variants for each inheritance pattern, it produces a spreadsheet with columns for “Sample Name”, “Inheritance Mode”, “Variant Rank”, “Score” (produced by the ABC), as well as the variant annotations from VCF. An analyst independently reviewed the ranked variants for each sample and determined which ranked variants were plausible solutions (Supplementary Tables 3, 4 and 6).

**Figure 2:**
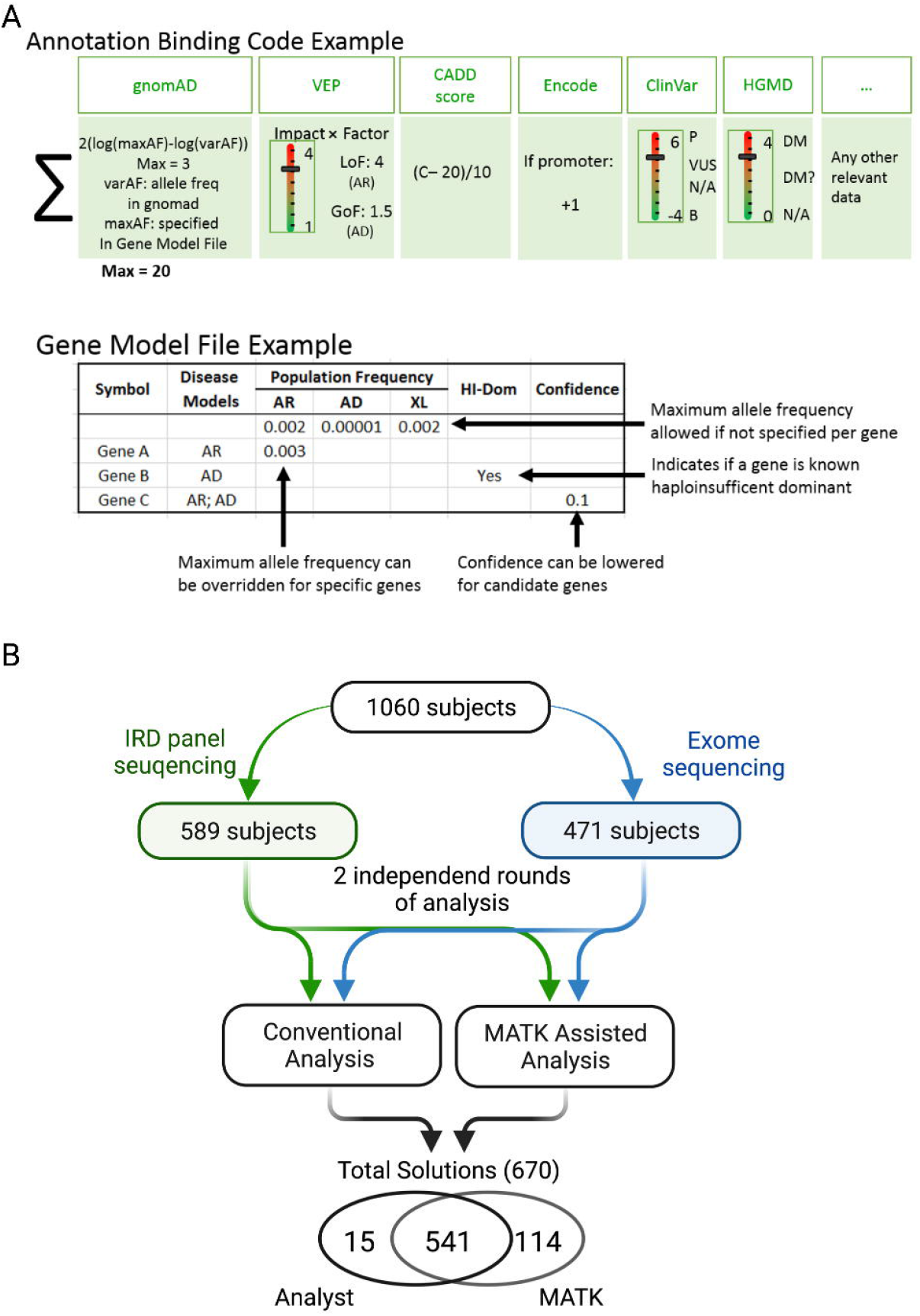
Functionality of MATK and study design. A) The two major components of MATK: The Annotation Binding Code (ABC), is the weighting function used to assign a score to each variant. The function used on the IRD cohort was tuned empirically, and utilized population frequency, sequence consequence, CADD scores, regulatory information, and prior reports, capped at 20 points. The Gene Model File (GMF) encapsulates disease specific gene-level information such as known inheritance modes, allele frequencies, haploinsufficient dominant status, and level of confidence in the field that a gene may be disease causing. B) 1060 IRD patients were analyzed with IRD-panel (589 subjects) and exome sequencing (471 subjects/families). Each sequence underwent two independent analysis rounds: with a conventional presentation of variants and hard filtering performed by the analysis and with MATK-assisted analysis. Solutions from both were analyzed for overlap and discrepancies.

For analysis of the ES cohort, a generalized version of MATK was used, without input of prior expert knowledge. The ABC was configured as a summation of 6 unique functions pertaining to 1) sequence consequence, 2) population frequency, 3) CADD score, 4) regulatory status 5) transcript expression level in retina^26^, and 6) variant quality score (all formulae are given in Supplementary Table 1B). The retina expression weighting function was calibrated using known IRD genes, where we determined that all IRD genes showed a superior expression of a lnTPM=2, where lnTPM is a natural log of gene expression represented as transcripts per million. We therefore added points to all genes lnTPM>2. The GMF was left empty aside from the default maximum likely frequencies of AR and XL at 0.001, and AD at 0.00001.

### Code availability

The software is open source, available at https://gitlab.com/matthew_maher/mendelanalysis

## RESULTS

### MATK functionality

The MATK software was developed to increase automation and accuracy of variant interpretation in the context of Mendelian disease. MATK is a suite of Python scripts that outputs a ranked list of variants and/or variant pairs according to a customizable set of rules and parameterized weighting functions, using sample-specific and gene-specific inputs. Sample-specific files include the annotated VCF, as well as optional inputs such as a structural variation file and pedigree information. Gene-specific files are meant to be customized for a given disease space and may include a curated list of known disease-associated genes with their expected causal variant frequencies and inheritance mode, or other gene-level information relevant to the studied disease (e.g. gene expression, pLI scores). Variant-level information is used in the Annotation Binding Code (ABC), a customizable component of the software that scores each variant for functional consequence, population frequency, ClinVar and/or HGMD pathogenicity classification, or *in silico* predictions of deleteriousness such as the CADD score. The ABC assigns scores differently to the genes with known or presumed autosomal recessive (AR), X-linked (XL) or autosomal dominant (AD) inheritance (Figure 2A). For example, loss-of-function variants are scored higher in AR, XL genes and AD haploinsufficient genes than in genes with AD inheritance known for a gain of function or dominant negative mechanism of disease. The gene-level information is encoded in the Gene Model File (GMF), which contains a default maximum likely frequency of a variant for a given disease or gene, which can be different for different inheritance modes, if the gene is known to contribute to both types of disease (Figure 2A, Supplementary Table 2). Once the GMF and ABC have executed, MATK calculates a score for each variant in a sample based on the specified inheritance mode. MATK then ranks the variants by score, and for the AR inheritance mode, creates ranked pairs of variants within each gene. The final step requires an analyst to determine if any of the ranked variants are accepted as candidate solutions. In this study we have measured MATK performance by a comparison of the MATK-assisted and conventional variant analysis of 1060 IRD subjects investigated with an IRD-specific gene panel (589 individuals) and exome sequencing (471 families) (Figure 2B).

### MATK-assisted analysis of panel sequencing data improves genetic diagnosis

To test the practicality of MATK, we performed a comparative analysis of a proband-only IRD subject cohort. We analyzed 589 primarily early-onset IRD samples in two independent rounds. The first round of analysis utilized our conventional analysis protocol, starting with an application of hard frequency and functional consequence filters to the dataset. The remaining variants are then evaluated based on frequency, variant type, computer prediction models, and prior reports to identify variants of interest. CNVs are analyzed separately, often second, with special attention to monoallelic (one potentially pathogenic variant in a recessive gene) cases or cases with no SNVs of interest. The second round of analysis incorporated the MATK software to prioritize the variants for analysts to review. For our cohort, we experimentally determined that displaying the first 20 variants for each inheritance mode was sufficient, although the number of MATK outputs can be adjusted based on preference. In both rounds, the analysts would choose one of two outcomes: the case was either potentially solved by a single or set of potentially pathogenic variants, or the case was left unsolved. Finally, all variants that were identified as potentially solving were classified using the ACMG/AMP guidelines, and based on these classifications, solutions were split into 3 confidence tiers: Tier 1 solutions comprised of likely pathogenic (LP) or pathogenic (P) variants; tier 2 solutions comprised of AR genes with one LP/P variant and one variant of uncertain significance (VUS); and tier 3 solutions comprised of two VUSs in an AR gene or one VUS in an AD gene. Samples in which only one variant in an AR gene was found and in which a chosen variant was classified as benign (B) or likely benign (LB) were considered as unsolved. Of the 589 cases, 502 (85.2%) had identical outcomes using both protocols, consisting of 323 potentially solved cases and 179 unsolved (Figure 3A, Supplementary Table 3). Overall, there were more solutions of each tier with the MATK-assisted analysis than in the conventional analysis (*χ*^*2*^ test, df=3, p=0.0005) (Figure 3A, B) and the MATK-assisted analysis identified 97.3% (323/332) of the potential solutions found by the conventional analysis. Of the 87 samples in which there was a discrepancy, 52 involved high confidence, tier 1 solutions. To further analyze the discrepancies, we considered only the 52 tier 1, high confidence solutions, because we believe that recording of the lower confidence solutions may be due to the analysts’ differences in stringency of variant interpretation rather than the analysis method. Forty-nine of the tier 1 discrepant solutions were discovered through the MATK assisted analysis and 3 via the conventional analysis (Figure 3A). Of the 49 tier 1 solutions found exclusively by MATK, 24 solutions involved structural variants, 16 solutions were missed in the conventional analysis due to human error and 8 were missed because of upstream pipeline errors related to issues with the variant caller, gene transcripts, and hard filters that resulted in the likely solving variants being hidden from the analysts. The last tier 1 solution missed in conventional analysis and found using MATK was a sample (OGI2423_003982) in which the two analysis protocols resulted in two different potential genetic solutions (Supplementary Table 3). In this case, the ACMG/AMP classification clarified that the MATK-assisted analysis chose the most likely solution. Of the three tier 1 solutions found exclusively using the conventional analysis, one solution was a mitochondrial solution (OGI2050_003476, NC_012920.1 MT-*ATP6* m.8993T>G), which the MATK program did not consider in any of its inheritance modes. One solution was missed because the gene transcript model incorrectly labeled a variant as non-coding, which caused the variant to be scored so low as to not appear in the MATK output (OGI2401_003960, NM_014249.3 *NR2E3*: c.932G>A, p.(Arg311Gln) ; c.151G>A, p.(Gly51Arg)). Finally, one solution (OGI1257_002429) was presented by MATK but was missed due to human error in interpreting a complex indel in *USH2A* (NM_206933.2: c.13335_13337del, c.13339A>T, c.13342_13347del), even though all data was available in MATK for the analyst to make the correct interpretation of c.13335_13347delinsCTTG, p.(Glu4445_Ser4449delinsAspLeu) (Supplementary Table 3).

**Figure 3:**
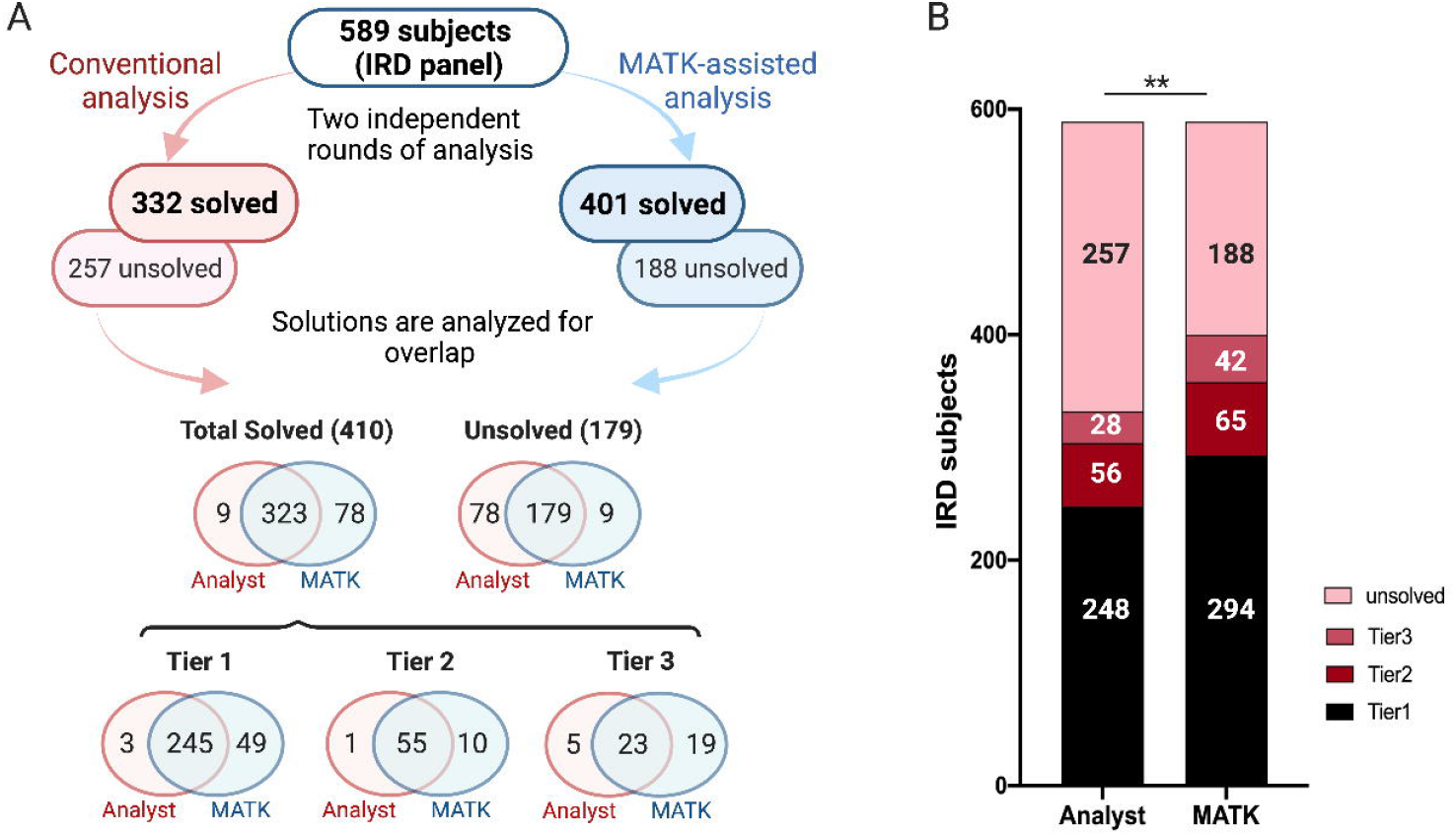
Comparison of 589 panel sequenced IRD patients analyzed with and without MATK assistance. A) A flow chart detailing all of the results obtained from the conventional and MATK-assisted variant analyses. There was a high degree of overlap between the two methods, with MATK assisted analysts missing only 9 potential solutions and non-MATK assisted analysts missing 78 potential solutions. The overlap of the solutions in all three confidence tiers is also presented. B) A bar graph illustrating the overall performance of both methods.

The remaining discrepant solutions (35 cases, 29 from the MATK assisted analysis and 6 from the conventional analysis) involved low confidence solutions (tier 2 and 3), which were interpreted differently in the two different analysis rounds. Reviewing these samples using the ACMG/AMP classification guidelines found that these potentially solving solutions had one or more VUS variants, and thus the genetic cause could not be resolved unequivocally. Therefore, for the purpose of the comparison between the conventional and MATK-assisted analysis, the 297 high-confidence solutions are most informative, where 294 (99.0%) were obtained with the MATK-assisted analysis, and 248 (83.5%) were obtained with the conventional analysis.

### MATK facilitates data reanalysis after upstream pipeline changes

Routine reanalysis of all the sequenced samples with a conventional protocol each time a reference genome, transcript model, other crucial database or software is updated is time consuming for a genetic diagnostic lab, however an automated process enables regular reanalysis. We have therefore used the MATK-assisted analysis to understand how updating the human reference genome and associated datasets can affect diagnostic rate in the known IRD genes. The sequence reads for all of 589 samples were realigned to hg38. Reanalysis with MATK and comparison with the existing solutions produced 13 additional potential solutions, four of which were ultimately placed in the tier 1 category. Of these four, one was found because of a new ClinVar entry that brought the variant to the attention of the analyst (OGI2484_004049, NM_001030311.2 *CERKL*: hom c.237_238+13del), one was found because of an updated transcript model (OGI2114_003548, NM_014249.3 *NR2E3*: hom c.932G>A, p.(Arg311Gln)), and two were found because of updated gCNV results (OGI2201_003682, NM_015629.3 *PRPF31*: hg38:chr19:54115160-54118353_duplication), and (OGI1943_003348, NM_015662.1 *IFT172*: hg38: chr2:27458072-27465609:deletion) (Supplementary Table 4).

### MATK improves accuracy of paired SNV and CNV calls

In three cases, where a heterozygous deletion paired with an overlapping pathogenic SNV, the variant was called as homozygous. The MATK-assisted analysis gave a more accurate genetic solution in each case. For sample OGI1820_003160, a nonsense variant in NM_006343.2 *MERTK* (hg19:chr2:112732995G>T, c.1090G>T, p.(Glu364Ter)) overlapped with a deletion of hg19:chr2:112655935-112787192. For OGI1973_003378, a known IRD pathogenic variant in NM_206933.2 *USH2A* (hg19:chr1: 216420460C>A, c.2276G>T, p.(Cys759Phe)) overlapped with a deletion of hg19:chr1:216380365-216424690. Lastly, for OGI2285_003796, a missense variant in NM_022787.3 *NMNAT1* (hg19:chr1: 10042553G>A, c.634G>A, p.(Val212Met)) overlapped with a deletion of hg19:chr1:10003159-10044256. (Supplemental Table 3).

### MATK comparison to Exomiser

We next compared MATK to Exomiser, an open source variant prioritization software^27^. For a subset of our panel sequenced cohort (96 samples), we ran Exomiser in PhenIX mode, which prioritizes variants in known human disease genes (Supplemental Methods). We then compared the Exomiser rankings to the previously established tier 1 solutions found by analysts using MATK. Data was reviewed to determine if Exomiser ranked the same solution, a “partial” solution (for example, ranking only one variant in an AR solution), or did not rank any of the established solution. Of 34 tier 1 solutions, Exomiser presented a full solution only for 17 cases (two variants in an AR and one variant in an AD or XL case) and in 11 cases only one variant from an AR solution was presented. Of the full solutions, 12 were presented in the top 3 suggestions and five in the top 25. A similar trend was observed in the tier 2 and 3 solutions presented by Exomiser. No additional solutions in the 39 unsolved IRD patients were revealed by the Exomiser analysis (Figure 4, Supplementary Table 5).

**Figure 4:**
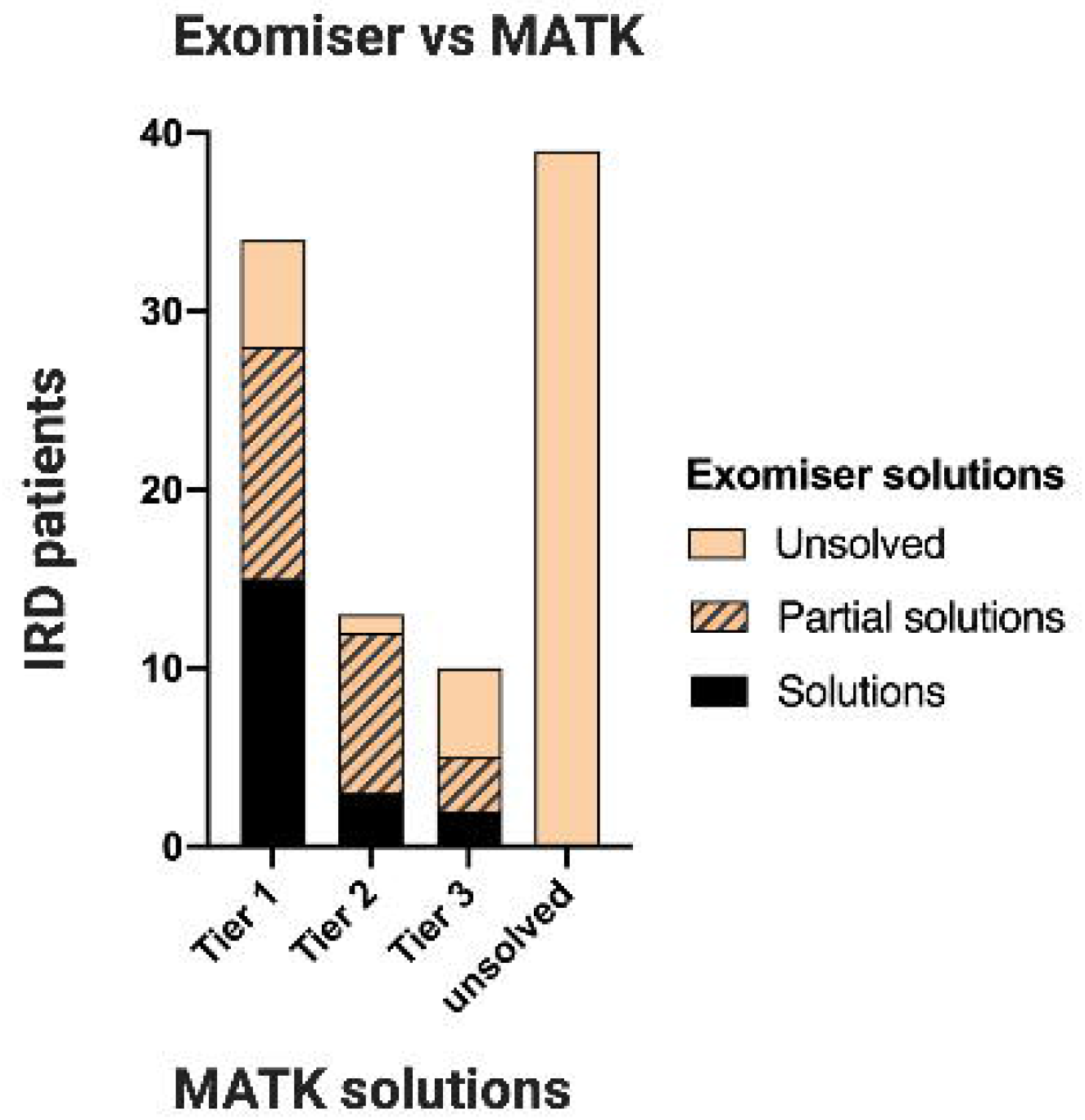
Comparison of Exomiser and MATK in 96 panel sequenced samples. Out of 56 total solutions found using MATK, Exomiser successfully ranked 20 total solutions, 15 in tier 1, three in tier 2, and two in tier 3, without finding any new solutions that were missed by MATK.

### MATK results in exome sequencing

To test the utility of MATK in the analysis of exome data (ES), we ran MATK on 471 exome sequenced cases, consisting of both proband-only cases and families. After examining the sequencing results with both the conventional analysis and MATK assisted analysis, we found the two protocols agreed in 429 (91.1%) of cases, consisting of 218 potentially solved (142 tier 1 samples, 39 tier 2 samples, 37 tier 3), and 211 unsolved cases (Figure 5A, B, Supplemental Table 6). The MATK-assisted analysis was able to find 97.3%, (218/224) of the solutions found by conventional analysis. Of the tier 1 solutions, there were 4 cases in which the conventional analysis found solutions that MATK missed. These consisted of one family with a partial penetrance inheritance pattern (OGI842, NM_015629.3 *PRPF31* c.73_166dup, p.(Asp56GlyfsTer33)), and three solutions that were found in a genomic region where difficulties in sequencing resulted in heterozygous calls on the X chromosome, even though the samples in question were male, and thus failed the inheritance checks in MATK (OGI1426, NM_001034853.1 *RPGR* c.1582_1585del, p.(Thr528LeufsTer4)), (OGI1741, NM_001034853.1 *RPGR* c.2909del, p.(Gly970GlufsTer119)), (OGI1735, NM_001034853.1 *RPGR* c.2506dup, p.(Glu836GlyfsTer243)). The MATK-assisted analysis found 26 possible new solutions missed by conventional analysis. Specifically, 20 of the 26 involved CNVs and 6 of the 26 were missed because of shortcomings in the analysis pipeline such as filtering out common pathogenic variants and gene transcript errors (Figure 5A, B, Supplementary Table 6).

**Figure 5:**
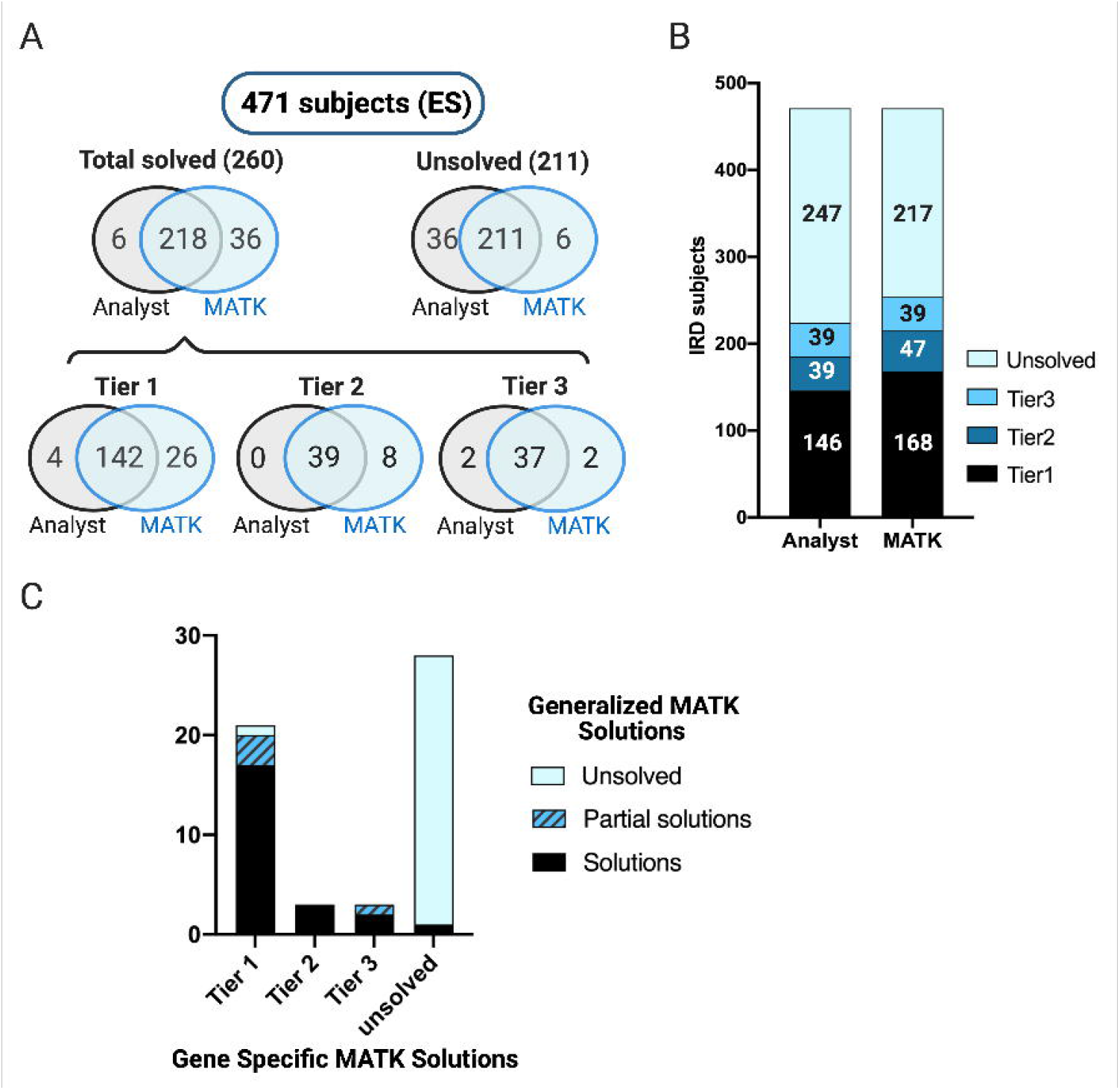
Comparison of 471 exome sequenced IRD patients analyzed with MATK versus the conventional analysis pipeline. A) Venn diagrams of the results obtained from the conventional and MATK-assisted variant analyses, showing a high degree of overlap between the two methods, with MATK assisted analysts missing only 6 potential solutions and non-MATK assisted analysts missing 36 potential solutions. B) A bar graph illustrating the overall performance of both methods C) Comparison of gene-specific MATK vs generalized MATK. Out of 27 total solutions, the generalized MATK successfully ranked 17 of 21 solutions in tier 1, with additional 3 partial solutions (monoallelic recessive solution). All tier 2 and 2/3 of tier 3 solutions were also fully identified. The generalized MATK was able to find one solution that was unsolved in the gene-specific MATK analysis run, but was determined to be a tier 1 solution.

Next, we tested a more generalized version of MATK which excluded the use of known IRD genes or prior variant reports. The purpose of this test was to produce a “mock scenario” in which MATK might be run on a disease cohort in an early gene discovery phase when less is known about genetic causality of that disease. For this trial, we took a subset of the ES cohort consisting of 55 families of trios or larger. As with Exomiser, we considered whether the generalized MATK ranked the established solutions fully, partially, or not at all. In order to narrow down the gene search without utilizing known disease genes or reported variants, we used publicly available gene expression data in the relevant tissue (in this case, retina)^26^ and used a weighting function that strongly discounted genes expressed at low levels in retina (see Methods). Of the 21 tier 1 solutions uncovered with the IRD-specific MATK, the generalized MATK recapitulated 17 fully, 3 partially, and missed one solution entirely. For tier 2 and tier 3 solutions, the generalized MATK performed similarly to the IRD-specific MATK, missing only one solution partially in each category (Figure 5C, Supplementary Table 6).

## DISCUSSION

We have demonstrated the utility of an automated variant analysis process in providing genetic diagnoses using a cohort of patients with retinal disorders. Our MATK software was practical and efficient for finding causal variants in the known IRD genes in targeted gene panel data and in ES data. The MATK-assisted analysis showed a higher number of plausible solutions than the non-MATK-assisted analysis, largely due to the customization options in MATK, which allow for incorporation of the existing scientific knowledge about the genetics of a disorder into the ranking algorithms. Even with minimal customization, many clear genetic solutions were still ranked highly by MATK in ES data. In addition, we showed the utility of MATK for rapid re-analysis of a large cohort after realignment to a new genome build and transcript model, which resulted in finding new solutions.

Comparison between the conventional and MATK-assisted analysis showed a high level of reproducibility, with the MATK-assisted analysis identifying 97.3% (541/556) of potential solutions that were found using a more conventional analysis in both IRD gene panel sequencing (323/332) and exome sequencing (218/224). Furthermore, the MATK-assisted analysis identified 78 additional potential solutions for the gene panel cohort, and 36 additional potential solutions in the exome cohort, the majority of which consisted of high confidence, tier 1 solutions (49/78 for the gene panel cohort and 26/36 for the exome cohort). Some of these new findings were easily anticipated, such as the solutions found with MATK’s improved utilization of the structural variation data. Although this discrepancy between MATK and conventional analysis should eventually be decreased by better incorporating CNV analysis into the conventional protocol, we believe that as more sophisticated annotations are developed, there will always be a lag in integrating them, which further highlights the advantage of using MATK to re-run variant ranking with new information. Some of the high confidence solutions were missed without MATK due to human error, which primarily involved samples with many rare variants passing the hard filtering thresholds, creating more visual noise in the variant viewer software used in the conventional analysis. However, some of the discrepancies exposed deeper shortcomings in our analysis pipelines, for example, certain issues in the variant annotation and filtering steps resulted in the solution not being available to the analysts. Since these upstream sequence data processing and annotation pipeline are often being updated and improved, we anticipate that regular data re-analysis will uncover such missing diagnoses. In the gene panel cohort, the MATK assisted analysis missed three high confidence solutions. Only one of these, the mitochondrial DNA solution in OGI2050_003476, represented a true shortcoming in the software. The other missed solutions erroneous variant annotation leading to inadequate variant scoring by the software, such as in the case of OGI2401_003960 where the gene transcript in *NR2E3* was labeled “noncoding”, which deprioritized the variant in the MATK algorithm. Or they were due to human error, such as in the case of OGI1257 _002429 where a complex indel in *USH2A* was not correctly identified as a pathogenic variant even though all data was available to the analyst. With the exome sequencing cohort, four high confidence solutions were missed in the MATK assisted analysis, all of which were due to atypical inheritance situations. This could be easily remedied in future runs of MATK by relaxing the inheritance requirements.

There were 35 discordant results in the gene panel cohort and 12 discordant results in the exome cohort where the potential solution involved at least one VUS (tier 2 and 3 solutions). We believe these discrepancies stem from the uncertainty, or differences in interpretation of the variant’s potential disease-involvement. Such discrepancies, even when using standardized guidelines, are common and have been reported previously^28^. Even though they are ambiguous, a potential recessive solution with one P/LP variant and one VUS can be worth further investigation, as shown in previous studies that determined a single pathogenic variant paired with a clear phenotypic association strongly implies the solution will be found in that gene^29^. Sequence data for a great number of subjects (113/589 for the gene panel cohort and 88/471 for the exome cohort) contain such ambiguous solutions. This further highlights the utility of using a software with a reproducible output to assist with variant analysis, as such VUS containing solutions should be periodically reinvestigated as more evidence is accumulated to push the variant toward either a pathogenic or benign interpretation.

Using MATK allowed us to identify three false homozygous SNV calls when a variant was most likely in trans with a heterozygous CNV deletion. Critically, the samples with false homozygous pathogenic calls would not have been further investigated in our conventional analysis protocol. This would lead to providing an inaccurate genetic diagnosis, incorrect genetic counseling to family members regarding their carrier status, and misreporting of the allele frequency of that variant. Not all subjects have family members available for segregation testing, however even in the case where segregation testing occurs, a false homozygous call may lead to an interpretation of non-paternity rather than CNV. In the era of emerging CRISPR-based genetic therapies, where variant or exon-specific therapies will one day be more common, such inaccurate diagnosis could impact the efficacy of a gene therapy^30^. For example, the ongoing clinical trial NCT03872479 involves a gene editing product specific for the intronic variant (*CEP290*: c.2991+1655A>G), and whether the patient is compound heterozygous versus homozygous could potentially reduce the benefit of the treatment.

For a subset of our patients, we have benchmarked MATK against another freely available variant prioritization software, Exomiser, using a setting designed for known human disease genes (PhenIX). For a well-described monogenic disease space such as IRDs, MATK outperforms Exomiser in finding likely pathogenic variants. Other software such as Variant Score Ranker^31^, Variant Ranker^32^, Diploid Moon, and EmedGene, have been developed in recent years to address the issue of variant prioritization and new gene discovery, and programs such as PathoMAN^14^ are designed to classify variants according to ACMG/AMP guidelines^13^. There has been great success in using such programs to help find disease causing variants^33,34^. Our MATK software differs from these in that it is far more customizable and can incorporate disease-specific information.

For IRDs, there are over 270 known disease genes^20^, and knowledge about the plausible pathogenic variant frequency and inheritance modes have been incorporated into the MATK via one of the input files, the Gene Model File. For such a thoroughly characterized field, a highly customized MATK helps to encode prior scientific knowledge. In our IRD panel test cohort, there were solutions found in 77 unique genes, and gene rarity varied widely, with *USH2A* accounting for 85 solutions, and 29 genes causing disease in a single individual in the cohort. This level of diversity can be demanding for an analyst to remain cognizant of. However, not all Mendelian disorders have been so well characterized and many new disease genes remain to be discovered. Thus, we have tested a more general configuration of MATK on an exome sequencing dataset. We emulated a search for new disease genes by removing the disease-specific genetic information and the use of prior genetic knowledge such as ClinVar scores, and by including retinal RNA expression data to identify genes that are relevant to eye tissue. For many disease spaces, other gene-level data sets could be included such as loss-of-function or missense intolerance (pLI) scores^7^, however these are not useful for IRDs because vision loss does not influence reproductive fitness and thus known pathogenic variants are not depleted in the general population as in other systemic diseases. Our results showed that MATK was able to prioritize most of the solutions and thus could be used as a first pass analysis for other Mendelian diseases with minimal customization. As more knowledge is acquired about a specific disease, MATK customization can be added in an iterative process, allowing all previously hard-won knowledge gains to accumulate to the benefit of future analysts. Even for such well characterized disorders as IRD, only about 60% of IRD patients receive genetic diagnoses in the known genes^21,22^, meaning there still may be undiscovered pathogenic genes. In future studies, especially as the field moves to sequence exomes and genomes of unsolved families, we would consider incorporating other dataset such as SpliceAI^35^, GO terms^36^, String terms^37^, HPO terms^38^, and Monarch data^39^ into MATK. Integrating well calibrated automated variant ranking tools into analysis protocols is a critical step in improving Mendelian disease diagnosis and new gene discovery in both clinical and research settings.

## Data Availability

Variants will be available through ClinVar (Submission Number SUB9430499, SUB9443633, and SUB9363246) and in Supplemental Materials. Sequence data will be available through dbGaP and upon request.

## Acknowledgments

This work was supported by grants from SPARK Therapeutics Inc. (EAP), the National Eye Institute [R01EY012910 (EAP), R01EY026904 (KMB/EAP) and P30EY014104 (MEEI core support)], and the Foundation Fighting Blindness [EGI-GE-1218-0753-UCSD, (KMB/EAP)]. Exome sequencing and analysis were provided by the Broad Institute of MIT and Harvard Center for Mendelian Genomics (Broad CMG) and was funded by the National Human Genome Research Institute, the National Eye Institute, and the National Heart, Lung and Blood Institute grant UM1HG008900 and in part by National Human Genome Research Institute grant R01 HG009141. The authors thank all subjects for their participation in this study and the OGI Genomics Core members for their experimental assistance.

## Author Information

Conceptualization: E.Z., M.M., E.M.P, K.M.B; Data curation E.Z., E.M.P., N.E.W., S.D., K.R.C., E.E, B.CMG, A.C., S.N., S.H., J.P., K.M.B.; Formal Analysis: E.Z., K.M.B; Funding acquisition: E.M.P., B.CMG, E.A.P., K.M.B.; Investigation: E.Z., M.M., B.CMG, E.G-B., K.M.B.; Software: M.M, C.F., B.C.; Visualization: E.Z., K.M.B.; Writing – original draft: E.Z, K.M.B; Writing – review & editing: E.Z, M.M., E.M.P., N.E.W., S.D., K.R.C., B.C., K.M.B., E.M.P.

## Notes

### Competing Interest Statement

The authors have declared no competing interest.

### Author Declarations

The study was approved by the Institutional Review Board at the Massachusetts Eye and Ear (Human Studies Committee MEE, Mass General Brigham, USA)

### Summary of Updates

This version of the manuscript was updated after peer review.

