## Supplemental Methods for "The Importance of Automation in Genetic Diagnosis: Lessons from Analyzing an Inherited Retinal Degeneration Cohort with the Mendelian Analysis Toolkit (MATK)" (from another version of this preprint: version 2, 2021-10-16)

Corresponding author:

Dr. Kinga Bujakowska

**Clinical evaluation**

Subjects included in the study were recruited and clinically examined at MEE. Ophthalmic examination included best-corrected Snellen visual acuity, dynamic Goldmann visual field testing, dark adaptation testing, and full-field electroretinogram (ERG) testing with assessment of 0.5 Hz ERG amplitude and 30 Hz ERG amplitudes.

**Sequencing and Annotation**

DNA was extracted from venous blood using the DNeasy Blood and Tissue Kit (Qiagen, Hilden, Germany). All samples underwent Genetic Eye Disease test (GEDi) sequencing as described previously^21^. For inclusion in the cohort, samples were required to have >20x depth of coverage for >90% of the reads. VCF files were generated using the Genome Analysis Toolkit (GATK) version 3 (https://software.broadinstitute.org/gatk/), and annotated using VEP^6^ and VCFAnno^23^. VEP provided transcript-specific sequence-consequence annotations from GENCODE v19 and regulatory annotations from ENCODE^24^. VCFAnno was used to annotate the variants with gnomAD^7,8^, ClinVar^10^, HGMD^11^, and CADD^9^. CNV predictions were produced using gCNV^4^, and the known *MAK*-Alu structural variant was identified using a custom script^25^.

Exome sequencing was performed at the Center for Mendelian Genomics at the Broad Institute of MIT and Harvard using methodology described previously^7^. Briefly, whole exome sequencing and data processing were performed by the Genomics Platform at the Broad Institute of MIT and Harvard with an Illumina Nextera or Twist exome capture (~38 Mb target) and sequenced (150 bp paired reads) to cover >80% of targets at 20x and a mean target coverage of >100x. Exome sequencing data was processed through a pipeline based on Picard and mapping done using the BWA aligner to the human genome build 38. Variants were called using Genome Analysis Toolkit (GATK) HaplotypeCaller package version 3.5. The data were displayed and analyzed with an online tool (https://seqr.broadinstitute.org).

**Exomiser Configuration**

Another variant ranking software, Exomiser^27^, was run using the code available at <https://exomiser.github.io/Exomiser/manual/7/example/test-analysis-genome/>. The PhenIX prioritizer and OMIM prioritizer were used. HPO terms used in the analysis were: ("HP:0000510", "HP:0000548", "HP:0007754", "HP:0000546", "HP:0000608", "HP:0007737", "HP:0007722", "HP:0008527", "HP:0008615", "HP:0030610", "HP:0030611"). Using the genePanelFilter option, we utilized the same gene list from the MATK GMF (Supplementary Table 2). The resulting scores were ordered first by EXOMISER_GENE_COMBINED_SCORE, then by EXOMISER_VARIANT_SCORE to create a ranking of variant scores.
